# Population-based cancer incidence and mortality rates and ratios among adults with intellectual disabilities in Scotland

**DOI:** 10.1101/2024.01.18.23300433

**Authors:** L. A. Ward, S-A. Cooper, F. Sosenko, D. Morrison, M. Fleming, C. McCowan, K. Robb, C. Hanna, L. Hughes-McCormack, K. Dunn, D. Conway, A. Henderson, G. Smith, M. Truesdale, D. Cairns

**Affiliations:** Health Informatics Centre, Division of Population and Health Genomics, School of Medicine, University of Dundee; School of Health and Wellbeing, Mental Health and Wellbeing, University of Glasgow; School of Medicine, Medical & Biological Sciences, University of St Andrews; School of Cancer Sciences, College of Medical Veterinary & Life Sciences, University of Glasgow; Glasgow Dental Hospital and School, School of Medicine, Dentistry & Nursing, University of Glasgow

**Keywords:** intellectual disabilities, cancer, health inequalities, data linkage

## Abstract

**Objective:** To provide contemporary data on cancer mortality rates within the context of incidence in the population with intellectual disabilities.

**Methods:** Scotland’s 2011 Census was used to identify adults with intellectual disabilities and controls with records linked to the Scottish Cancer Registry and death certificate data (March 2011-December 2019). The control cohort without intellectual disabilities and/or autism were used for indirect standardisation and calculation of Crude Incident Rates/Crude Mortality Rates (CIR/CMR), and age-sex Standardized Incident Rate Ratios/ Standardized Mortality Ratios (SIR/SMR), with 95% Confidence Intervals (CI).

**Results:** Adults with intellectual disabilities were most likely diagnosed cancers of digestive, specifically colorectal (14.2%), lung (9.3%), breast (female 22.9%), body of the uterus (female 9.3%) and male genital organs (male 17.6%). Higher incident cancers included metastatic cancer of unknown primary origin (female SIR=1.70, male SIR=2.08), body of uterus (female SIR=1.63), ovarian (female SIR=1.59), kidney (female SIR=1.85), and testicular (male SIR=2.49). SMRs were higher, regardless of a higher, similar, or lower incidence (female SMR=1.34, male SMR=1.07). Excess mortality risk was found for colorectal (male SMR=1.59), kidney (female SMR=2.85u), female genital organs (ovarian SMR=2.86u, body of uterus SMR=2.11), breast (female SMR=1.58), and metastatic cancer of unknown primary origin (female SMR=2.50u, male SMR=2.84).

**Conclusions:** Adults with intellectual disabilities were more likely to die of cancer than the general population. Reasons for this may include later presentation/diagnosis (so poorer outcomes), poorer treatment/compliance, or both. Accessible public health approaches are important for people with intellectual disabilities, and healthcare professionals need to be aware of the different cancer experiences faced by this population.

**Summary box:** Strengths and limitations of this study

- Our key strength is the comprehensive coverage of Scotland’s entire adult population with intellectual disabilities, and inclusion of a representative general population comparison group.
- By using nationwide robust data linkage of high-quality electronic health records, we provide reliable data with minimal bias.
- Limitations include our inability to account for cancer incidence before the census date. However, prospective collection of data over nearly 9 years provided well-powered person-time for rate calculation, allowing for a meaningful interpretation of mortality rates in the context of incidence.
- Death certificate data imprecision is considered, but our dual-analysis (main-cause and all-cause analyses), mitigates differences and indeed have similar interpretations.

## 1. Introduction

Intellectual Disabilities are a group of conditions with significant limitations in intellectual functioning and adaptive behaviour, with onset in childhood affecting 1.4% of the world population (1). People with intellectual disabilities continue to face substantial health inequalities culminating in a 20-year premature mortality gap (2), and a higher proportion of avoidable deaths compared with the general population (3,4). One of the most common avoidable mortalities is cancer, as many cancers are considered either preventable or treatable (5). However, there lacks robust comparatives studies of cancer incidence and mortality between the population with and without intellectual disabilities. This is crucial, as healthcare assumptions based on general population evidence may not be applicable for the population with intellectual disabilities.

Cancer is a leading cause of mortality (6), but studies about people with intellectual disabilities show inconsistent findings. Cohort studies indicate a higher Standardised Mortality Ratio (SMR) for people with intellectual disabilities compared to the general population (4,7,8), though some report no significant difference (9). The most common cancer-related deaths in the population with intellectual disabilities include respiratory (lung), digestive (colon), and breast cancers (7,10). Disparities compared to the general population were highest for digestive, metastatic cancer with unknown primary origin, bladder, and cervical cancers (SMRs between 2-3); lip, oral cavity, and pharynx, rectal, female genital organs, colon, oesophageal, haematopoietic, urinary, breast and pancreatic cancers (SMRs between 1-2). Cuypers and colleagues found no cancers associated with a lower mortality rate (7). Specific data on stomach, liver, body of uterus, ovarian, testicular, kidney or brain cancers were not reported in this extensive work. Higher rates of colorectal cancer mortality have been reported in males (SMR=2.7), but not females with intellectual disabilities (9). Glover and colleagues reported that women with intellectual disabilities had greater risk of female genital organ cancer mortality (SMR=2·3), however this was based on nine deaths split between cancer of the body of the uterus and ovary, with exact figures not reported (9). Unlike Cuypers et al., non-significant SMRs for breast, lung, and haematopoietic cancers were reported, and authors agreed that brain cancer was not significantly different in this population (9). A smaller study found no statistical differences in mortality from breast, lung, and digestive cancers in adults with intellectual disabilities, perhaps due to the study size (11). These conflicting results (with wide confidence intervals, different cancer categories, and varying age ranges), highlights the gaps in the scientific literature available on cancer mortality ratios in the population with intellectual disabilities.

Cancer mortality is the combination of cancer incidence (being diagnosed with cancer), survival rates and the occurrence of both cancer-related death and non-cancer-related death. Therefore, whether the reported higher SMRs are due to a higher incidence, later presentation, or poorer care is yet to be determined. People with intellectual disabilities have distinctly different factors that could influence likelihood of cancer incidence; for example, a higher prevalence of obesity, gastroesophageal reflux disorder, exposure to helicobacter pylori infection, more sedentary behaviour and mobility problems, poorer diets, and nulliparity, but a lower likelihood to smoke or drink alcohol excessively (12). National studies consistently evidence lower screening program participation in the population with intellectual disabilities (13), potentially impacting cancer rates.

The incidence of cancer in the population with intellectual disabilities appears to be lower in older adults (14,15), higher in children and young adults (16), or the same as in the general population for children and adults combined (17,18). However, methodological limitations exist in this evidence base, including retrospective study design (excluding people with incident cancer who died), inclusion of people with autism (who may have different health profiles), and sampling cohorts from those using support services or hospital discharge records (14–16). Similarly, identification via residential care received identified <35% of people with intellectual disabilities reported in Cuyper and colleagues’ mortality paper which used more extensive methods (19), and the authors confirm this likely focuses on people with more severe intellectual disabilities (7). Older studies from Patja and Sullivan and colleagues used more robust identification methods, and are better comparators for our results, despite identifying only 70% of the population via service-use (17,18). However, the data are more than 20 years old (1967-1997; 1982-1997), so may not reflect more recent cancer rates due to lifestyle changes (namely long-stay hospital closure and community care for adults with intellectual disabilities). However, the population with intellectual disabilities had higher incident cancers of gall bladder and thyroid cancers, and lower prostate and lung cancers (17). Leukaemia, corpus uteri and colorectal cancers were reported as higher in females with intellectual disabilities, leukaemia, brain, and stomach cancers were more common in males with intellectual disabilities, whilst prostate cancer was less common (18). Each of these studies report a similar wide range of common cancer types, and except for a lower incidence of prostate cancer, the findings are contradictory.

The aim of this study was to describe both cancer incidence and mortality rates in people with intellectual disabilities at a population level using a large, nationwide cohort of adults of all ages with intellectual disabilities, compared with the general population.

## 1. Methodology

### 2.1 Data Sources and Study Population

Population data from Scotland’s 2011 Census linked to the National Records of Scotland (NRS) death certificate data and Scottish Cancer Registry (Scottish Morbidity Records 06, SMR06) held by National Services Scotland were used. As previously described (20), linkage was undertaken for the 94% of the Scottish population who completed Scotland’s 2011 Census. The cohorts consisted of all adults with intellectual disabilities (with or without co-occurring autism aged 18+) as recorded within the Census, and a 15% randomly selected comparator sample from the general population who had neither intellectual disabilities nor autism. Record linkage between census and health records was successful for >92% of these two cohorts. We report cancer incidence and mortality for an 8-year, 9-month period from 28/03/2011 (1 day after Scotland’s 2011 Census) to 31/12/2019 (prior to excess Covid-19 mortality and under-recorded cancer incidence). Cases who were alive at the end of the study were censored on the study end date. This study excluded 30 cases from the general population who did not self-identify as having an intellectual disability in the Census record, but subsequently died during the study with one or more all-contributing factors relating to an intellectual disability or autism (ICD-10 codes F70, F71, F72, F73, F78, F79, F84). Self-reported data on biological sex at birth were taken from the Census but there were a small number with mismatched sex-specific cancers, e.g., females with prostate cancer. These individuals were included in overall cancer rates but excluded from sex-specific cancer rates. Although there were proportionately more mismatched sex cases in the population with intellectual disabilities compared to the general population, the number of mismatches in the former was so small (<5) that excluding mismatched records did not have any meaningful impact on either rates or ratios. The number of records in the linked analysis dataset was 583,264.

### 2.2 Data Variables and Management

Baseline demographics of age, sex, Scottish Index of Multiple Deprivation (SIMD 2016) quintile and living arrangements (intellectual disabilities group only) were taken from Scotland’s 2011 Census (https://www.scotlandscensus.gov.uk/about/2011-census/). SIMD is a composite measure derived from geographical area of residence relating to socioeconomic status (https://www.gov.scot/collections/scottish-index-of-multiple-deprivation-2020). NRS Death records were used to identify details of deaths, with the main cause of death defined internationally as the ‘disease or injury which initiated the chain of morbid events leading directly to death’. Given concerns about the quality of recording on death certificate data on main cause of death, like others we chose also to combine the all-contributing causes of death (up to 10 additional causes) with cancer mentioned on the death certificate in any position. Presented cancer mortality results are those people who have died with cancer as main cause of death listed in position 1; all-cause mortality results are reported in the Supplementary Tables (S4, S5).

The Cancer Registry includes information on all new diagnoses of cancer occurring within Scotland. NRS death data and Cancer Registry include diagnostic codes from the International Classification of Disease 10th revision (ICD-10) and specific cancers were grouped accordingly (S1). The Cancer Registry holds data on tumour types using the International Classification of Diseases for Oncology (ICD-O). Scotland’s Cancer Registry has high quality robust population data; however, there is a necessary time delay to allow accrual of information. In the whole cohort there were <0.5% discrepancies between the Cancer Registry and death certificate data, and data from the Cancer Registry were prioritized, e.g., NRS death data coding for ‘Colon unspecified cancer’ and Cancer registry coding for ‘Rectal cancer’. However, NRS deaths are updated daily and there were <1.0% cases of cancer-related mortality without available data from the Cancer Registry matching death certificate data.

### 2.3 Statistical Analysis

Baseline characteristics were reported from the time of Scotland’s 2011 Census. Cancer incidence and mortality rates were calculated from any newly diagnosed cancers (incidence) and cancer-related deaths (mortality) during the study period. Numbers reported are for cancer per person, with percentages calculated from the total number of cancers not person, as individuals with multiple cancers are included in different categories, e.g., lung and breast cancer. Crude Incidence and Mortality Rates (CIR/CMR) are reported per 100,000 person-years using the cancer diagnosis date/ date of death and reflect the cancer burden faced by each group separately. Age-sex-Standardized Incidence Rate Ratios and Mortality Ratios (SIR/SMR) are reported with the general population as reference (indirectly standardised), with 95% Confidence Intervals (CI). Ratios (SIR/ SMR) higher than 1.0 indicate an increased risk for the population with intellectual disabilities, and less than 1.0, a lower risk. Rates are reported per 100,000 person-years unless there are fewer than <5 cases where no calculation was attempted due to lack of reliability and cases are reported as <5 due to disclosure risk. Totals above 5 are similarly suppressed where providing the exact total would disclose the number below 5 for a specific sex in further tables (i.e., a suppression of <10 and <20 is also used). For cancer types that figures have been suppressed, percentages are not reported. All rates calculated from variables within 5-20 deaths are labelled as unreliable (“u”) in line with the Office for National Statistics guidelines (5). Non-melanoma skin cancer (ICD-10 code C44) was excluded from statistical analyses of all cancers combined, due to incomplete incidence data capture, and in line with Public Health Scotland guidelines (21). One researcher (LW) conducted the analyses, and a second researcher (FS) verified the coding for accuracy. All statistical analysis was conducted in Stata version 16.

#### Ethical considerations

The research was approved by Scotland’s Public Benefit and Privacy Panel for Health (1819-0051), Scotland’s Statistics Public Benefit and Privacy Panel (1819-0051), and the University of Glasgow’s College of Medical, Veterinary, and Life Sciences Ethical Committee (200180081). The first author affirms that the manuscript is an honest, accurate, and transparent account of the project. All summary results are available within the manuscript or supplementary files and no important aspects have been omitted. Funding for the study includes the UK Medical Research council (grant MC_PC_17217), the Scottish Government via the Scottish Learning Disabilities Observatory (SLDO), and the Baily Thomas Charitable Fund. The SLDO has patient and public involvement in the steering committee where people with intellectual disabilities, carers and public members can guide, review, and disseminate all research conducted.

## 3. Results

### 3.1 Baseline characteristics of study cohort

The linked datasets consisted of 17,203 adults (136,590 person-years) with intellectual disabilities (with/without autism) and 566,061 adults (4,683,379 person-years) from the general population without intellectual disabilities or autism. As expected, the intellectual disabilities cohort contained a greater number of males, were on average younger and resided in more deprived areas. Table 1 shows the baseline characteristics of the whole cohort taken at the time of Scotland’s 2011 Census and the cohort with any cancer diagnosis or cancer-related death. There were 3,240 (18.8%) adults with intellectual disabilities who died during the study, 435 (2.5%) were cancer-related mortalities, compared to 64,339 (11.4%) deaths in the general population with 18,678 (3.3%) cancer-related deaths. Figure 1 shows the different age structures between the groups of adults with and without intellectual disabilities, and the cohorts with cancer. The population with intellectual disabilities is a younger cohort due to the prevailing 20-year premature mortality health inequality, and this should be considered in the context of cancers, which are mostly age-related diseases.

**Table 1.**
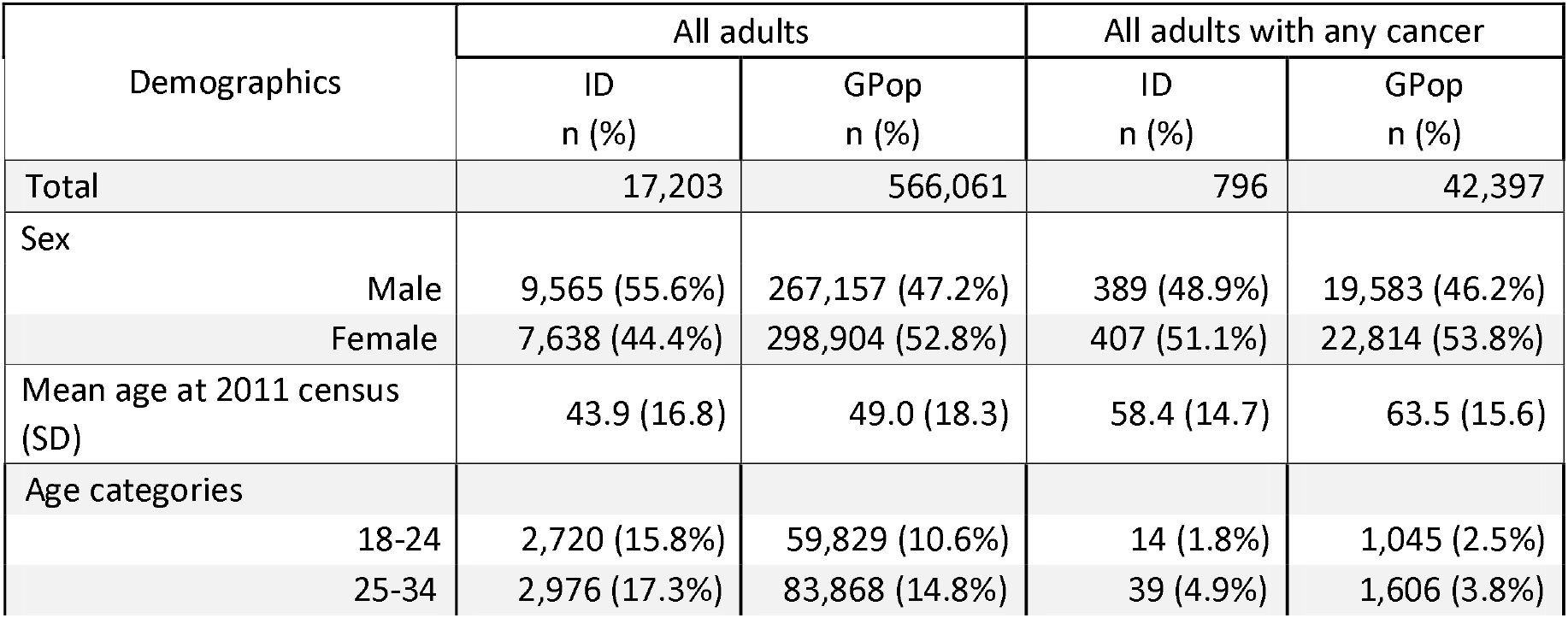
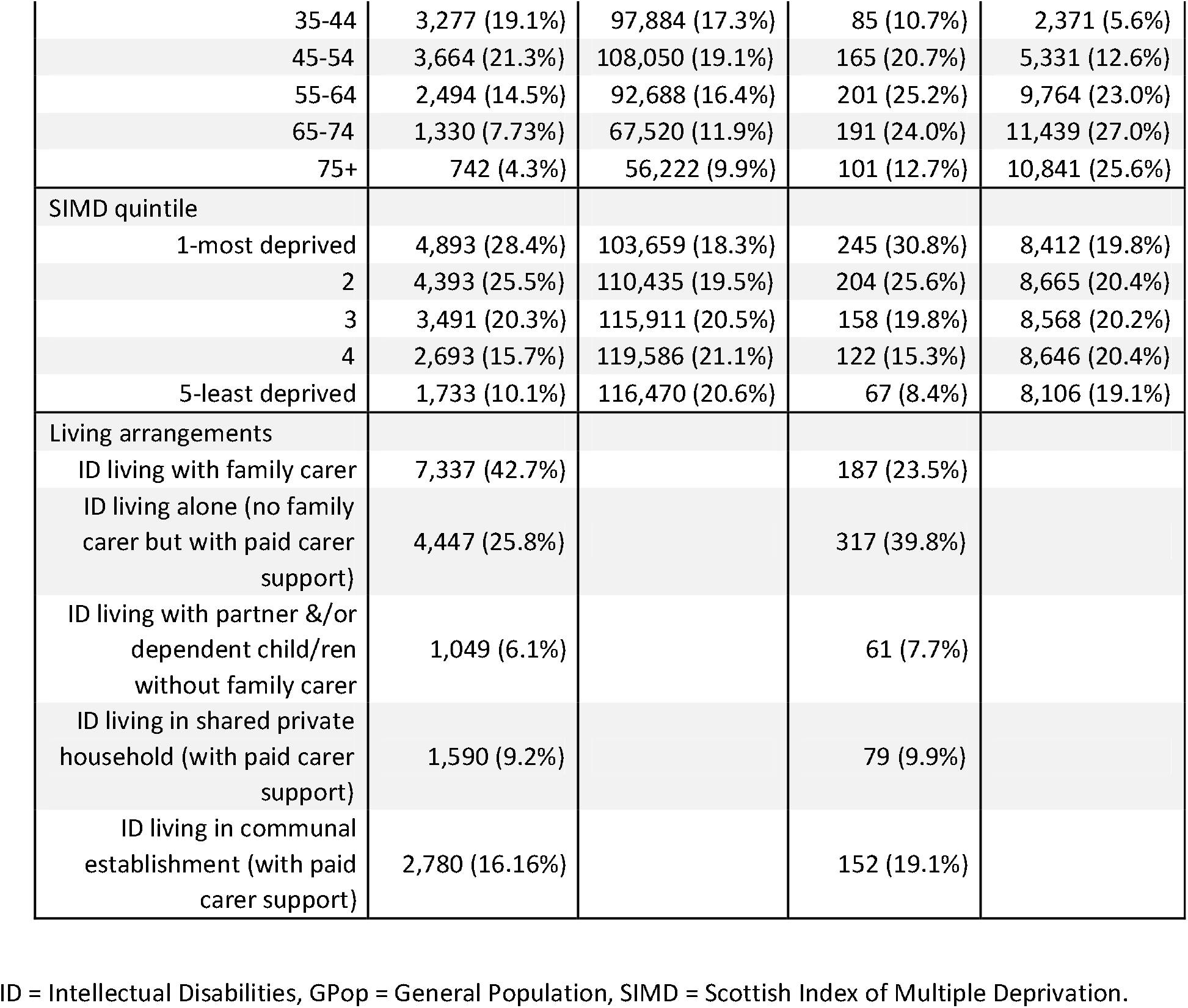
Demographic information for the whole cohort of adults (aged 18+, n=583,264) with and without Intellectual Disabilities (ID) and those with cancer diagnosis and/or cancer-related death (n=43,193).

| Demographics | All adults |  | All adults with any cancer |  |
| --- | --- | --- | --- | --- |
|  | ID<br>n (%) | GPop<br>n (%) | ID<br>n (%) | GPop<br>n (%) |
| Total | 17,203 | 566,061 | 796 | 42,397 |
| Sex |  |  |  |  |
| Male | 9,565 (55.6%) | 267,157 (47.2%) | 389 (48.9%) | 19,583 (46.2%) |
| Female | 7,638 (44.4%) | 298,904 (52.8%) | 407 (51.1%) | 22,814 (53.8%) |
| Mean age at 2011 census<br>(SD) | 43.9 (16.8) | 49.0 (18.3) | 58.4 (14.7) | 63.5 (15.6) |
| Age categories |  |  |  |  |
| 18-24 | 2,720 (15.8%) | 59,829 (10.6%) | 14 (1.8%) | 1,045 (2.5%) |
| 25-34 | 2,976 (17.3%) | 83,868 (14.8%) | 39 (4.9%) | 1,606 (3.8%) |
| 35-44 | 3,277 (19.1%) | 97,884 (17.3%) | 85 (10.7%) | 2,371 (5.6%) |
| 45-54 | 3,664 (21.3%) | 108,050 (19.1%) | 165 (20.7%) | 5,331 (12.6%) |
| 55-64 | 2,494 (14.5%) | 92,688 (16.4%) | 201 (25.2%) | 9,764 (23.0%) |
| 65-74 | 1,330 (7.73%) | 67,520 (11.9%) | 191 (24.0%) | 11,439 (27.0%) |
| 75+ | 742 (4.3%) | 56,222 (9.9%) | 101 (12.7%) | 10,841 (25.6%) |
| SIMD quintile |  |  |  |  |
| 1-most deprived | 4,893 (28.4%) | 103,659 (18.3%) | 245 (30.8%) | 8,412 (19.8%) |
| 2 | 4,393 (25.5%) | 110,435 (19.5%) | 204 (25.6%) | 8,665 (20.4%) |
| 3 | 3,491 (20.3%) | 115,911 (20.5%) | 158 (19.8%) | 8,568 (20.2%) |
| 4 | 2,693 (15.7%) | 119,586 (21.1%) | 122 (15.3%) | 8,646 (20.4%) |
| 5-least deprived | 1,733 (10.1%) | 116,470 (20.6%) | 67 (8.4%) | 8,106 (19.1%) |
| Living arrangements |  |  |  |  |
| ID living with family carer | 7,337 (42.7%) |  | 187 (23.5%) |  |
| ID living alone (no family carer but with paid carer support) | 4,447 (25.8%) |  | 317 (39.8%) |  |
| ID living with partner &/or dependent child/ren without family carer | 1,049 (6.1%) |  | 61 (7.7%) |  |
| ID living in shared private household (with paid carer support) | 1,590 (9.2%) |  | 79 (9.9%) |  |
| ID living in communal establishment (with paid carer support) | 2,780 (16.16%) |  | 152 (19.1%) |  |

ID = Intellectual Disabilities, GPop = General Population, SIMD = Scottish Index of Multiple Deprivation.

**Figure 1.**
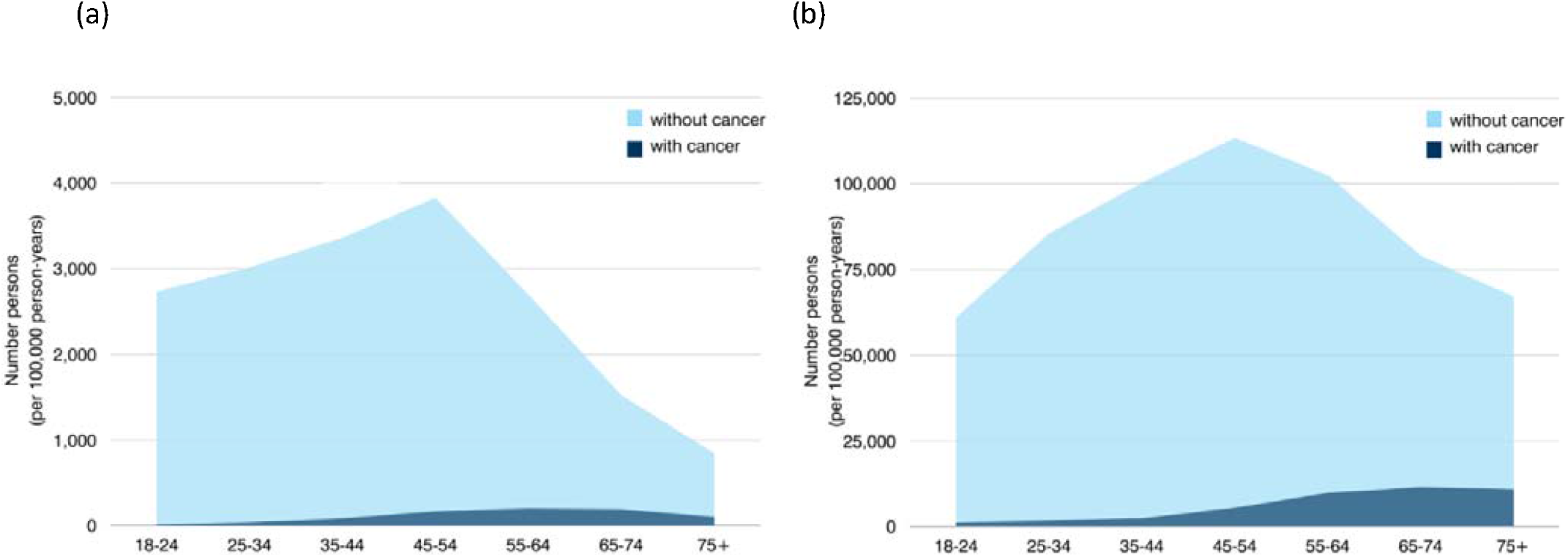
Age structure of cohorts with number of people per 100,000 person-years and incidence of any cancer by group for (a) adults with intellectual disabilities and (b) general population, number of people categorized into age groups form Scotland’s 2011 Census.

### 3.2 Cancer Incidence

Fewer adults with intellectual disabilities had a record of cancer (816 cancers in 796 people out of the 17,203 persons [4.7%]), compared with adults from the general population (43,775 cancers in 42,397 people out of the 566,061 persons [7.7%]). Table 2 shows cancer incidence aggregated by sex, Standardized Incident Rate Ratio for total SIR=0.76 (0.70-0.82), female SIR=0.79 [0.71-0.88], male SIR=0.71 [0.64-0.80] (total incidence is shown in S2). For women with intellectual disabilities, the most common diagnosis was breast (22.9% of all cancers among women), digestive (20.3%), specifically colorectal (10.0%) and female genital organs (17.2%), specifically body of the uterus (9.3%). For men with intellectual disabilities, the most common diagnosis was digestive (34.0% of all cancers among men), specifically colorectal (18.6%), male genital organs (17.6%), specifically prostate cancer (8.8%), and respiratory organs (12.8%). Additionally, 12 (2.9%) people with, and 2,290 (9.9%) without intellectual disabilities had cervical carcinoma-in-situ diagnosed.

**Table 2.**
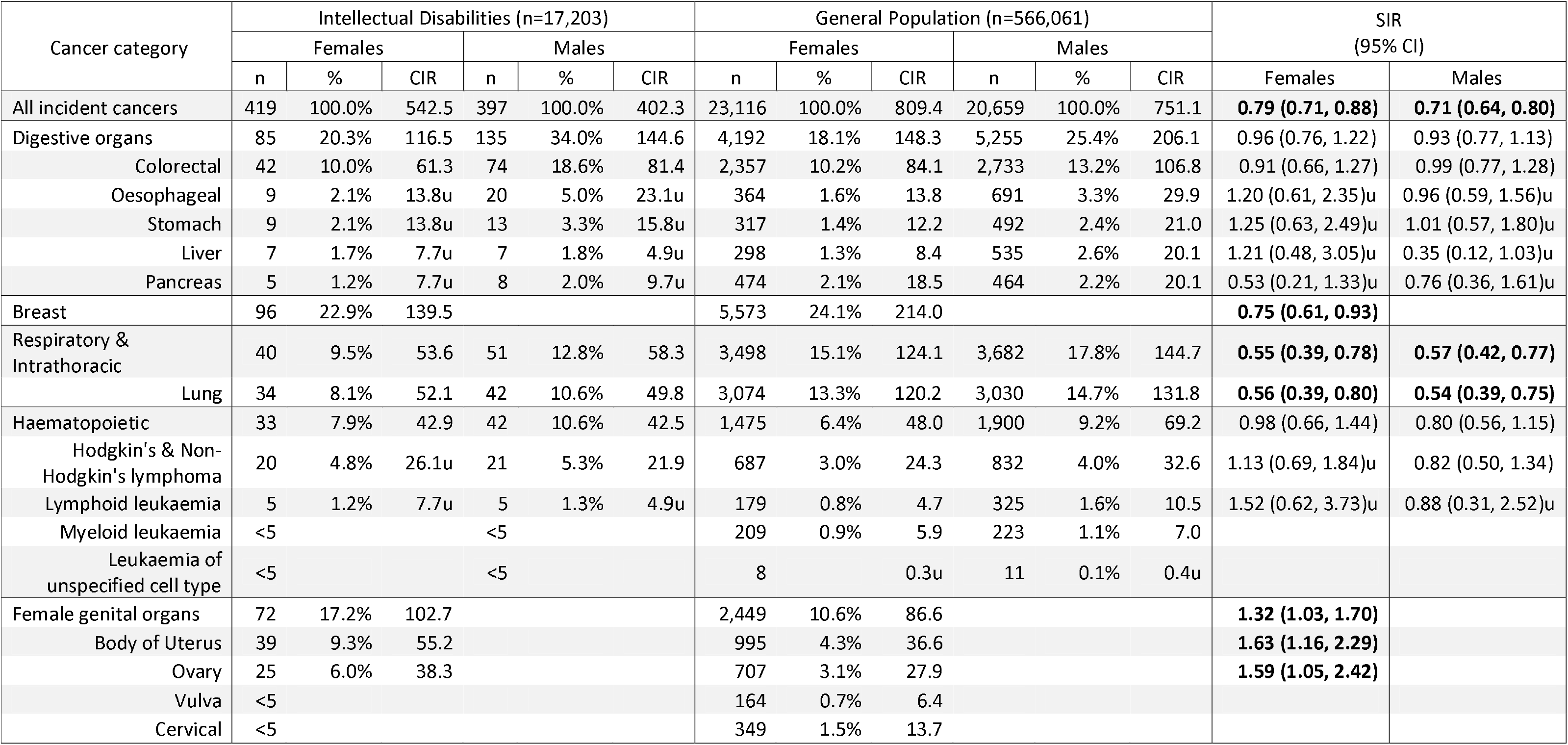
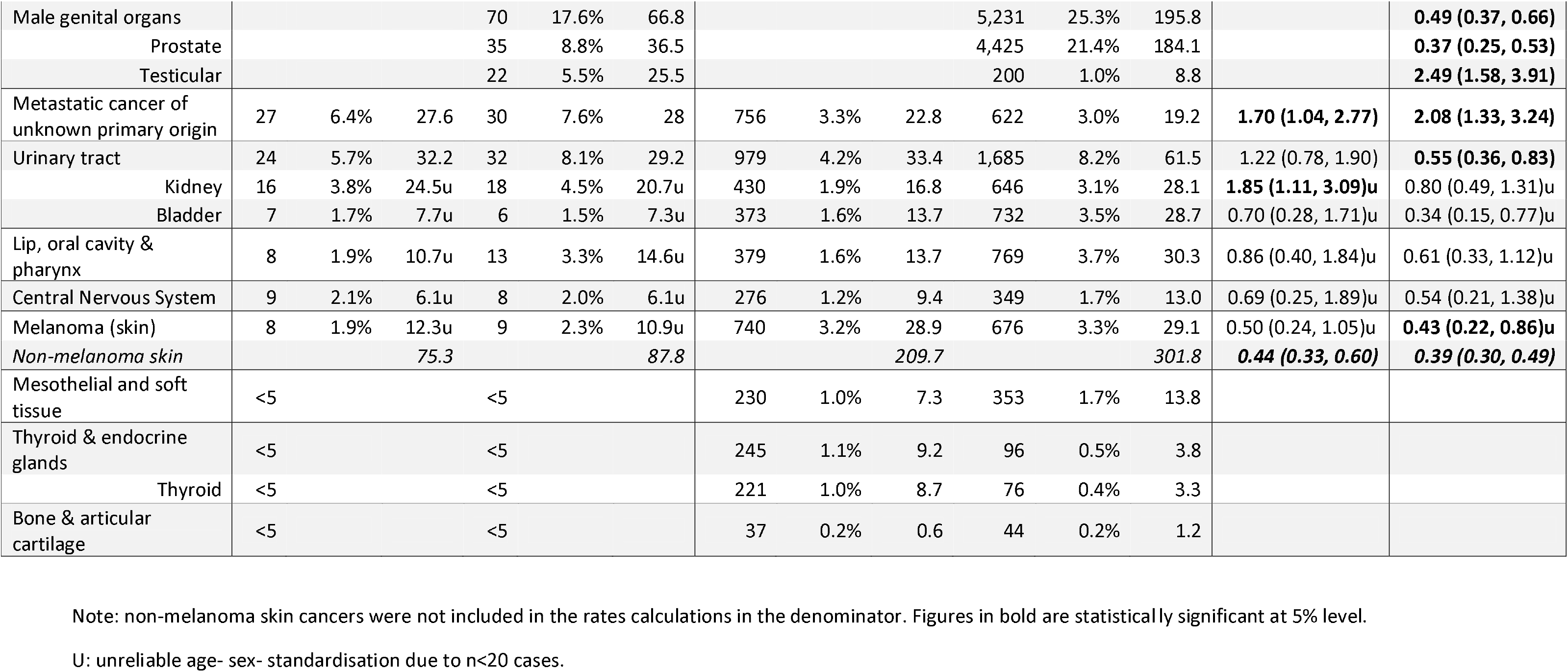
Cumulative cancer incidence by sex (raw numbers [n], percentages [%], Crude Incidence Rate [CIR], and Standardized Incidence Rate Ratio [SIR] with 95% Confidence Interval [CI]). Numbers reported are for cancer incidence, with percentages calculated from the total number of cancers not person. CIR are reported per 100,000 person-years and SIR are age-standardised rate ratios.

### 3.3 Cancer Mortality

During follow-up, cancer was the main cause of death for 435 (2.5%) of the 17,203 adults with intellectual disabilities and 18,678 (3.3%) of the 566,061 adults from the general population; SMR=1.20 (1.08, 1.33), female SMR=1.34 (1.16, 1.55), male SMR=1.07 (0.92, 1.24). Table 3 shows cancer mortality aggregated by sex, and total mortality is shown in S3.

**Table 3.**
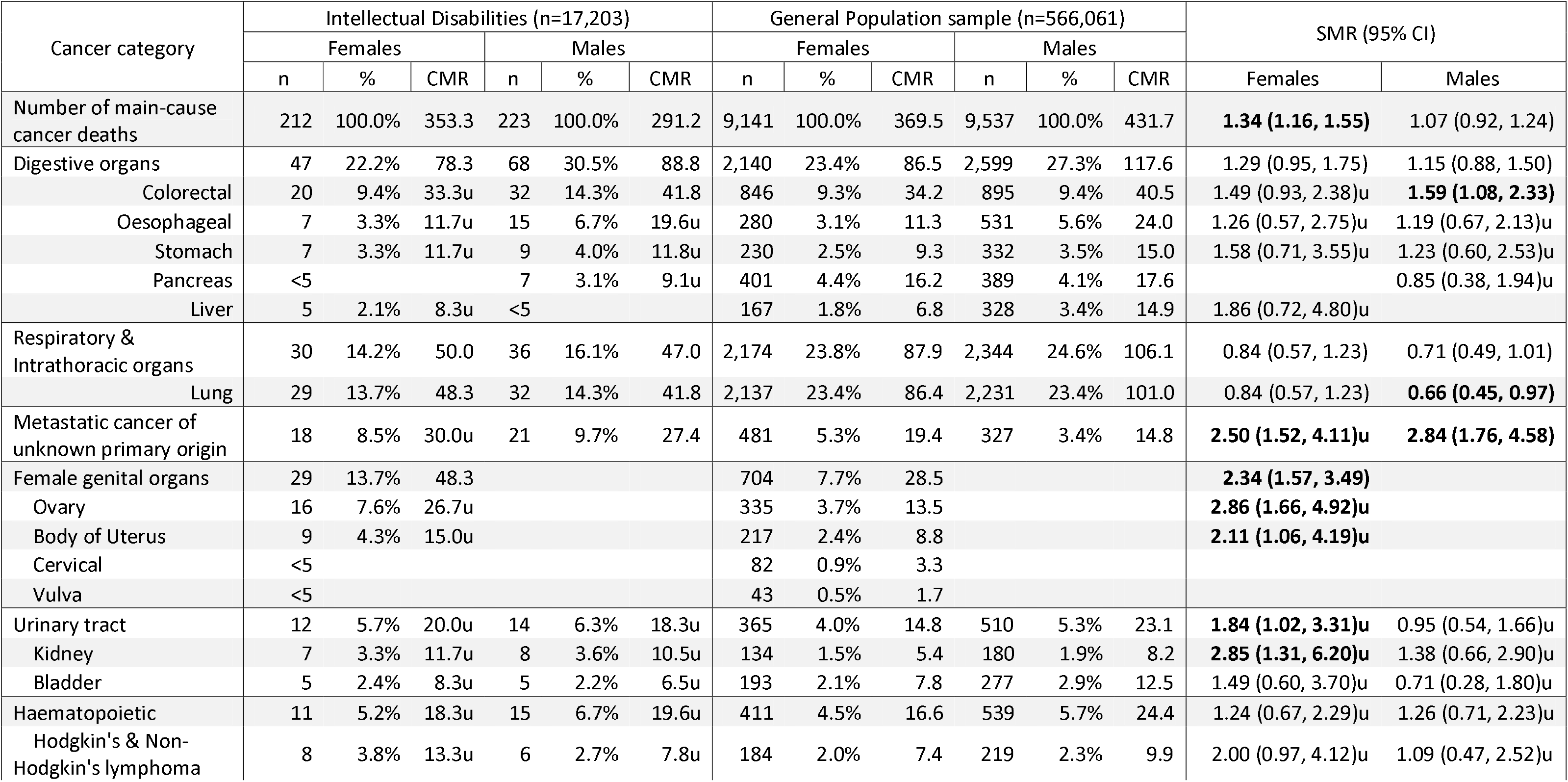
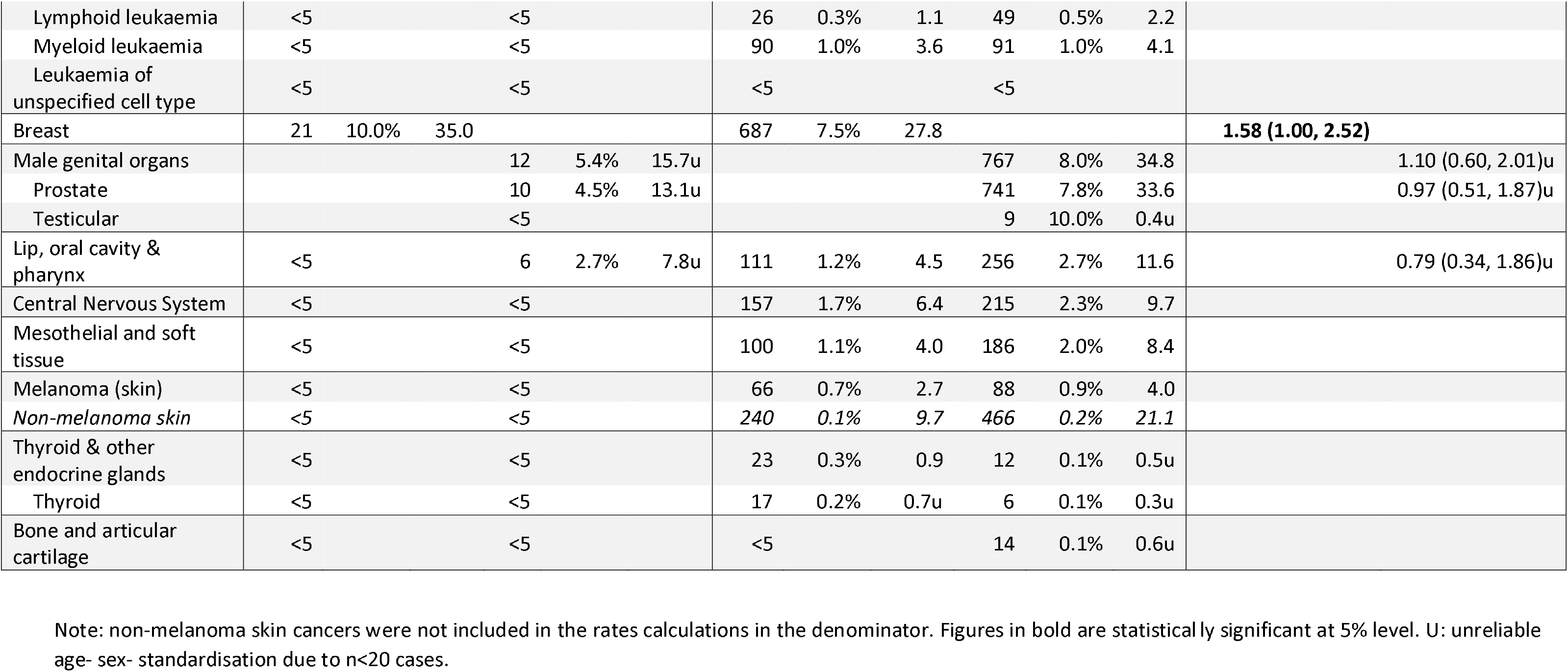
Cancer mortality by sex (raw numbers [n], percentages [%], Crude Mortality Rate [CMR], Standardized Mortality Ratios [SMR] with 95% Confidence Intervals [CI]), for main cause of death as cancer. CMR are reported per 100,000 person-years and SIR are age-standardised rate ratios.

Figure 2 plots trends in cancer incidence and mortality together with the most common cancers for each group, by sex. Standardised rate ratios comparing the population with and without intellectual disabilities show a clear trend of higher cancer deaths despite a lower or comparable incidence. For females this is most notable for cancers of the female genital organs (SIR=1.32, SMR=2.34), specifically ovarian (SIR=1.59, SMR=2.86u), body of uterus cancers (SIR=1.63, SMR=2.11u), and breast cancer (SIR=0.75, SMR=1.58). For males, the disparity between incidence and mortality is highest for colorectal cancers (SMR=1.59), and haematopoietic cancers (SIR=0.80, SMR=1.26).

**Figure 2.**
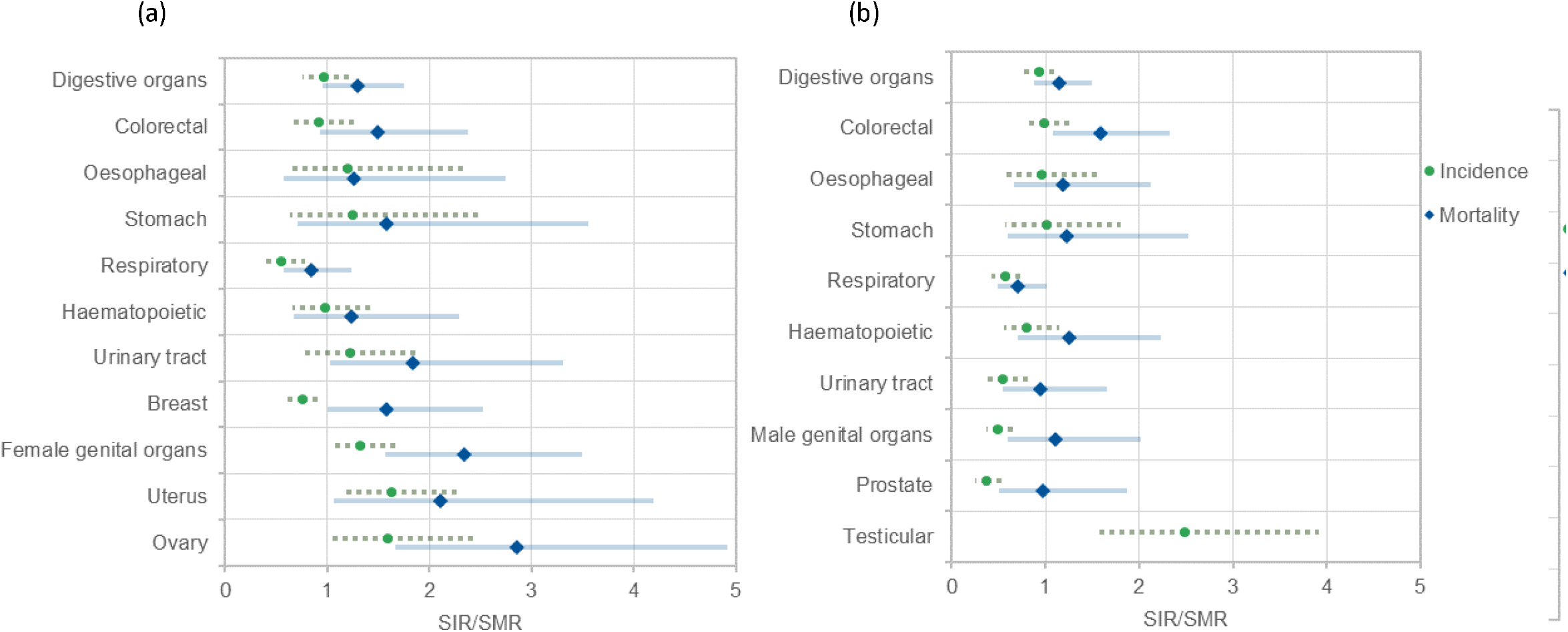
Age-sex-standardised rate ratios of commonest cancer incidence and mortality comparing adults with and without intellectual disabilities for (a) Females and (b) Males. Note there is no result for testicular mortality ratio due to fewer than <5 cases and no calculation attempted due to lack of reliability. Additionally, metastatic cancers of unknown primary origin are not included in these figures as cancer categories are primary cancers.

## 4. Discussion

The population with intellectual disabilities had higher cancer mortality rates than incidence rates across all cancer types, indicating poorer outcomes. This is the first study to report cancer-related mortality within the context of cancer incidence comparing adults with and without intellectual disabilities. We report excess mortality regardless of whether the number of diagnosed cancers were lower, higher, or comparable to the general population. Particularly striking was our finding that metastatic cancer of unknown primary origin had significantly higher incidence (SIR=1.86) and mortality (SMR=2.64) in the population with intellectual disabilities, demonstrating later presentation of cancer compared to the general population, potentially indicating a delay to diagnosis.

In terms of the most common cancer types, there are similarities and differences from the general population. Women with intellectual disabilities share the top two cancers with the general population (breast and colorectal), with female genital organ cancer being the third. Their incidence was higher for ovarian cancer (SIR=1.59), body of the uterus (SIR=1.63), ovarian (SIR=1.59), and kidney cancer (SIR=1.85u); and lower for breast cancer (SIR=0.75). Men with and without intellectual disabilities shared common cancers; digestive (specifically colorectal), male genital organs, and respiratory cancers, but with variations in testicular (SIR=2.49), and prostate cancer rates (SIR=0.37). The limited available evidence from older comparator studies has some support for higher incident digestive and uterine cancers in people with intellectual disabilities (18), but not for metastatic cancers of unknown primary origin. However, recent evidence indicates that adults with intellectual disabilities have more advanced cancer at diagnosis and poorer survival (22). This study reported a higher likelihood of preventable secondary cancers in people with intellectual disabilities; breast and colorectal, indicating that like our data, people with intellectual disabilities present later with cancer. However, this cross-sectional study reported a high rate of missing data (e.g., 33% of staging data for lung cancer in the intellectual disabilities, double that for the general population), and potential limitations in case identification of those with intellectual disabilities. Notably, our study reveals significantly higher rates of ovarian cancer in women with intellectual disabilities, a unique finding not previously reported (17,18). Similarly, in women with and without intellectual disabilities, comparable rates of breast cancer have previously been reported (17,18), but we found a statistically significant lower SIR. Breast cancer screening is crucial to avoid a proportion of breast cancer deaths through early treatment (10,23). However, in Scotland, women with intellectual disabilities were 45% less likely to participate in mammography screening, which may have contributed to lower detection rates (13). For men with intellectual disabilities, our results confirm lower prostate cancer incidence (17,18), and, for the first time, report higher testicular cancer incidence. While other cancers in our data could be compared to Patja and Sullivan, differences in study context and size should be noted, as well as the inclusion of children in these earlier studies.

Adults with intellectual disabilities were less likely to be diagnosed with cancer but were disproportionately more likely to die from cancer (SMR=1.20). The observed mortality rates were consistently elevated compared to the expected rates. Similar findings were reported by Cuypers and colleagues (SMR=1.48), although their analysis included in-situ and benign cancers we intentionally excluded (7). Common cancer-related deaths were similar for women with and without intellectual disabilities, including digestive (specifically colorectal), respiratory, and breast cancers. However, women with intellectual disabilities had higher mortality rates from female genital organ cancers (ovarian cancer SMR=2.86u, body of uterus SMR=2.11u), cancers of unknown primary origin (SMR=2.50u), and breast cancer (SMR=1.58). Common cancer-related deaths in men with intellectual disabilities mirrored the general population, including digestive (specifically colorectal), and respiratory cancers. However, men with intellectual disabilities experienced excess mortality from cancers of unknown primary origin (SMR=2.84) and colorectal cancer (SMR=1.59). These results confirm Cuypers et al. findings who report excess mortality for female genital organs (SMR=1.7), breast (SMR=1.43), digestive (SMR=1.59), and cancers of unknown primary origin (SMR=2.48). However, they did not report SMRs for kidney, ovarian, and uterine cancers, which we observed to be higher in the population with intellectual disabilities (7). Although our colorectal rate (SMR=1.54) is lower than previously reported (between 1.24-2.56) (7,9), there is a clear need to increase bowel screening participation for people with intellectual disabilities. Data from the Learning Disabilities Mortality Review (LeDeR) showed that 43% of people with intellectual disabilities who died with colorectal cancer were below the age threshold for screening (<60 years), suggesting a need to adjust public health programs for this population (23). This data suggests that public health approaches and messaging around breast, colorectal, and lung cancers are important for the population with intellectual disabilities (as well as for everyone else) and need to be accessible. Metastatic cancer of unknown primary origin results underscore the necessity for early detection and improved management of cancer. Factors contributing to higher SIRs and SMRs are likely complex, including self-care challenges, reliance on support workers to recognise cancer symptoms and signs, communication barriers, navigating health care services, and inexperience of many healthcare workers working with adults with intellectual disabilities.

Two noteworthy points are our differences in respiratory and cervical mortality rates compared to Cuypers et al. Our lower lung cancer mortality rate (SMR=0.75) contrasts their SMR=1.24 (7), but crude rates are similar (CMRs of 53 and 44.7). This suggests a lower smoking rate in the population with intellectual disabilities, but that the differing SMR directions is due to the lower general population rates in the Netherlands compared to Scotland. For cervical cancer, Cuypers et al., report a high SMR=1.94 (with 17 deaths), whereas our small numbers were potentially disclosive. Our cervical carcinoma-in-situ data suggest that the rarity is not due to screening, with only 12 women with intellectual disabilities diagnosed. This may be related to reduced sexual activity, potentially lowering HPV infection rates and cervical cancer. Whilst some women with intellectual disabilities may require support for cervical screening, our findings indicate other cancers contribute more significantly to excess cancer-related deaths. Assumptions about contributory behavioural and modifiable factors cannot be generalised between the population with and without intellectual disabilities. Indications in the data, such as living arrangement patterns, suggest the importance of exploring these factors in future research. Survival analyses are also indicated for common cancer types, as are studies on cancer staging at the time of presentation, cancer treatments, and compliance. Such information is crucial for improving cancer outcomes in this population.

Our study’s key strengths lie in the inclusion of a representative general population comparison group, and comprehensive coverage of Scotland’s entire adult population with intellectual disabilities (both living in private households and communal establishments). This population is difficult to identify in administrative health datasets, and the use of Scotland’s 2011 Census (with a high coverage rate) allows for self-identification. Robust record linkage also enhances data reliability and minimises bias. Prospective collection of data over nearly nine years, provided ample person-time for statistically well-powered analyses, allowing for a meaningful interpretation of mortality rates in the context of incidence. Limitations include our inability to account for cancer incidence before the census date. Non-melanoma skin cancer incidence may be undercounted due to registry data-capture issues in these cancers, though mortality data are comprehensive. Death certificate data imprecision is possible, given multiple clinicians completion, but our dual-analysis approach mitigates differences and indeed both have similar interpretations. Despite using national data, low absolute case numbers for some cancers limit the study’s power to detect differences.

## Conclusions

Patterns of cancer incidence and mortality differ between adults with and without intellectual disabilities. Public health strategies must consider the unique needs of people with intellectual disabilities, emphasizing accessibility. Promoting awareness of cancer symptoms among carers is crucial, especially for early detection. Support for bowel and breast screening programs is essential, addressing lower uptake rates observed in this population. While cervical screening is provided, its impact on reducing cancer deaths may be limited. Clinicians need to be aware that cancers can present late in this population and provide preventive interventions on known risk factors to reduce incidence.

## 5. Statements

### Author contributions

LW: conceptualization, methodology, formal analysis, data curation, writing - original draft, project administration.

S-AC: conceptualization, writing - original draft, writing – reviewing and editing.

FS: conceptualization, methodology, formal analysis, data curation, writing – reviewing and editing.

DM: conceptualization, methodology, formal analysis, writing – reviewing and editing.

MF: methodology, formal analysis, writing – reviewing and editing.

CM: methodology, writing – reviewing and editing.

KR: conceptualization, writing – reviewing and editing.

CH: methodology, writing – reviewing and editing.

LH-M: data curation, writing – reviewing and editing.

KD: writing – reviewing and editing.

AH: project administration, writing – reviewing and editing.

GS: methodology, writing – reviewing and editing.

MT: supervision, writing – reviewing and editing.

DC: conceptualization, supervision, writing – reviewing and editing.

## Acknowledgements

The authors would like to acknowledge the support of Public Health Scotland’s electronic Data Research and Innovation Service (eDRIS) team involved in the provisioning and linkage of the data, and for the use of the Trusted Research Environment for analysis within the National Safe Haven.

## Data availability statement

Data used in this study were available in the Scottish National Safe Haven (Project Number: 1819-0051), but as restrictions apply, they are not publicly available. Access to data may be granted on application to, and subject to approval by, the Public Benefit and Privacy Panel for Health and Social Care. Applications are coordinated by eDRIS (electronic Data Research and Innovation Service). The anonymised data used in this study was made available to accredited researchers only through the Public Health Scotland (PHS) eDRIS User agreement.

