## Supplementary Tables for "Population-based cancer incidence and mortality rates and ratios among adults with intellectual disabilities in Scotland"

S1. Cancer types categorised by ICD-10 codes from C00 – C97 Malignant Neoplasms.

| ICD10 code category | ICD10 subcategory | Cancer area |
| --- | --- | --- |
| C00-C14 |  | Lip, oral cavity, & pharynx |
| C15-C26 |  | Digestive organs |
|  | C15 | Oesophageal |
|  | C16 | Stomach |
|  | C22 | Liver |
|  | C18-C21  C25 | Colorectal  Pancreas |
| C30-C39 |  | Respiratory & Intrathoracic organs |
|  | C33-34 | Lung cancer |
| C40-C41 |  | Bone & articular cartilage |
| C43-C44 | C43  C44 | Melanoma (skin)  Other malignant neoplasms (non-melanoma) |
| C45-C49 |  | Mesothelial & soft tissue |
| C50 |  | Breast |
| C51-C58 | C51 | Female genital organs  Vulva |
|  | C53 | Cervical |
|  | C54-C55  C56 | Uterus  Ovary |
| C60-C63 |  | Male genital organs |
|  | C61 | Prostate |
|  | C62 | Testicular |
| C64-C68 | C64 | Urinary tract  Kidney |
|  | C67 | Bladder |
| C69-C72 |  | Eye, brain & other parts of the CNS |
| C73-C75 |  | Thyroid and other endocrine glands |
|  | C73 | Thyroid |
| C76-C80 |  | Metastatic cancer of unknown primary origin (ill-defined, secondary, & unspecified sites) |
| C81-C96 |  | Stated or presumed to be primary, of lymphoid, haematopoietic, and related tissue |
|  | C81 - C85 | Hodgkin’s & Non-Hodgkin’s lymphoma |
|  | C91  C92  C96 | Lymphoid leukaemia  Myeloid leukaemia  Leukaemia of unspecified cell type |

S2. Cumulative cancer incidence (raw numbers [n], percentages [%], Crude Incidence Rate [CIR], and Standardized Incidence Rate Ratio [SIR with 95% Confidence Interval [CI]). Numbers reported are for cancer incidence, with percentages calculated from the total number of cancers not person. CIR are reported per 100,000 person-years and SIR are age- sex- standardised rate ratios.

| Cancer category | Intellectual Disabilities (n=17,203) | | | General Population (n=566,061) | | | SIR  (95% CI) |
| --- | --- | --- | --- | --- | --- | --- | --- |
|  | n | % | CIR | n | % | CIR |  |
| All incident cancers | 816* | 100.0% | 464.3 | 43,775* | 100.0% | 781.8 | **0.76 (0.70, 0.82)** |
| Digestive organs | 220 | 26.9% | 132.2 | 9,447 | 21.5% | 175.6 | 0.95 (0.81, 1.10) |
| Colorectal | 116 | 14.2% | 72.5 | 5,090 | 11.6% | 94.8 | 0.96 (0.78, 1.17) |
| Oesophageal | 29 | 3.5% | 19 | 1,055 | 2.4% | 21.4 | 1.04 (0.70, 1.55) |
| Stomach | 22 | 2.7% | 14.9 | 809 | 1.8% | 16.4 | 1.11 (0.71, 1.72) |
| Liver | 14 | 1.7% | 6.1u | 833 | 1.9% | 13.9 | 0.63 (0.31, 1.27)u |
| Pancreas | 13 | 1.6% | 8.8u | 938 | 2.1% | 19.3 | 0.64 (0.36, 1.15)u |
| Respiratory & Intrathoracic | 91 | 11.1% | 56.3 | 7,180 | 16.4% | 133.8 | **0.56 (0.45, 0.70)** |
| Lung | 76 | 9.3% | 50.8 | 6,104 | 13.9% | 125.7 | **0.55 (0.43, 0.70)** |
| Haematopoietic | 75 | 9.2% | 42.7 | 3,375 | 7.7% | 58.0 | 0.88 (0.68, 1.14) |
| Hodgkin's & Non-Hodgkin's lymphoma | 41 | 5.0% | 23.7 | 1,519 | 3.5% | 28.2 | 0.96 (0.68, 1.36) |
| Lymphoid leukaemia | 10 | 1.2% | 6.1u | 504 | 1.1% | 7.4 | 1.09 (0.54, 2.20)u |
| Myeloid leukaemia | 5 | 0.6% | 2.7u | 432 | 1.0% | 6.4 | 0.62 (0.21, 1.80)u |
| Leukaemia of unspecified cell type | <5 | 0.0% |  | 19 | 0.0% | 0.4u |  |
| Metastatic cancer of unknown primary origin | 57 | 7.0% | 27.8 | 1,378 | 3.1% | 21.1 | **1.86 (1.34, 2.60)** |
| Urinary tract | 56 | 6.8% | 30.5 | 2,664 | 6.1% | 46.7 | 0.80 (0.59, 1.09) |
| Kidney | 34 | 4.2% | 22.4 | 1,076 | 2.5% | 22.2 | 1.22 (0.85, 1.76) |
| Bladder | 13 | 1.6% | 7.5u | 1,105 | 2.5% | 20.8 | 0.46 (0.25, 0.85)u |
| Lip, oral cavity & pharynx | 21 | 2.6% | 12.9 | 1,148 | 2.6% | 21.6 | 0.69 (0.43, 1.11) |
| Central Nervous System | 17 | 2.1% | 6.1u | 625 | 1.4% | 11.1 | 0.60u (0.30, 1.21)u |
| Melanoma (skin) | 17 | 2.1% | 11.52u | 1,416 | 3.2% | 29.0 | **0.47 (0.28, 0.78)u** |
| *Non-melanoma skin cancer* |  |  | *82.3* |  |  | *253.1* | ***0.41 (0.34, 0.50)*** |
| Mesothelial and soft tissue | 6 | 0.7% | 3.4u | 583 | 1.3% | 10.4 | 0.38 (0.15, 0.99)u |
| Thyroid & endocrine glands | 5 | 0.6% | 3.4u | 341 | 0.8% | 6.7 | 0.60 (0.24, 1.48)u |
| Thyroid | <5 |  |  | 297 | 0.7% | 6.1 |  |
| Bone & articular cartilage | <5 |  |  | 81 | 0.2% | 0.9 |  |

* This figure excludes non-melanoma skin cancers. Figures in bold are statistically significant at 5% level.

U: unreliable age- sex- standardisation due to n<20 cases.

S3. Cancer mortality (raw numbers [n], percentages [%], Crude Mortality Rate [CMR], Standardized Mortality Ratios [SMR] with 95% Confidence Intervals [CI]), for main cause of death as cancer. CMR are reported per 100,000 person-years and SMR are age- sex- standardised rate ratios.

| Cancer category | Intellectual Disabilities (n=17,203) | | | General Population (n=566,061) | | | SMR  (95% CI) |
| --- | --- | --- | --- | --- | --- | --- | --- |
|  | n | % | CMR | n | % | CMR |  |
| Number of main-cause cancer deaths | 435 | 100.0% | 318.5 | 18,678 | 100.0% | 398.8 | **1.20 (1.08, 1.33)** |
| Digestive organs | 115 | 26.4% | 84.2 | 4,739 | 25.4% | 101.2 | 1.21 (0.99, 1.48) |
| Colorectal | 52 | 12.0% | 38.1 | 1,741 | 9.3% | 37.2 | **1.54 (1.13, 2.08)** |
| Oesophageal | 22 | 5.1% | 16.1 | 811 | 4.3% | 17.3 | 1.22 (0.76, 1.93) |
| Stomach | 16 | 3.7% | 11.7u | 562 | 3.0% | 12.0 | 1.38 (0.80, 2.35)u |
| Pancreas | <20 |  | 8.1u | 790 | 4.2% | 16.9 | 0.74 (0.39, 1.42)u |
| Liver | <10 |  | 5.1u | 495 | 2.7% | 10.6 | 0.76 (0.34, 1.72)u |
| Respiratory & Intrathoracic organs | 66 | 15.2% | 48.3 | 4,518 | 24.2% | 96.5 | **0.77 (0.59, 1.00)** |
| Lung | 61 | 14.0% | 44.7 | 4,368 | 23.4% | 93.3 | **0.75 (0.57, 0.98)** |
| Metastatic cancer of unknown primary origin | 39 | 9.0% | 28.6 | 808 | 4.3% | 17.3 | **2.64 (1.86, 3.74)** |
| Urinary tract | 26 | 6.0% | 19.0 | 875 | 4.7% | 18.7 | 1.32 (0.87, 2.00) |
| Kidney | 15 | 3.4% | 11.0u | 314 | 1.7% | 6.7 | **2.01 (1.16, 3.49)u** |
| Bladder | 10 | 2.3% | 7.3u | 470 | 2.5% | 10.0 | 1.03 (0.54, 1.99)u |
| Haematopoietic | 26 | 6.0% | 19.0 | 950 | 5.1% | 20.3 | 1.25 (0.82, 1.90) |
| Hodgkin's & Non-Hodgkin's lymphoma | 14 | 3.2% | 10.3u | 403 | 2.2% | 8.6 | 1.51 (0.87, 2.60)u |
| Lymphoid leukaemia | <5 |  |  | 75 | 0.4% | 1.6 |  |
| Myeloid leukaemia | <5 |  |  | 181 | 1.0% | 3.9 |  |
| Leukaemia of unspecified cell type | <5 |  |  | <5 |  |  |  |
| Lip, oral cavity & pharynx | <20 |  | 6.6u | 367 | 2.0% | 7.8 | 0.96 (0.48, 1.95)u |
| Central Nervous System | 7 | 1.6% | 5.1u | 372 | 2.0% | 7.9 | 0.76 (0.34, 1.70)u |
| Mesothelial and soft tissue | <5 |  |  | 286 | 1.5% | 6.1 |  |
| Melanoma (skin) | <5 |  |  | 154 | 0.8% | 3.3 |  |
| *Non-melanoma skin* | *<5* |  |  | *706* | *0.1%* | *15.1* |  |
| Thyroid & other endocrine glands | <5 |  |  | 35 | 0.2% | 0.8 |  |
| Thyroid | <5 |  |  | 23 | 0.1% | 0.5 |  |
| Bone & articular cartilage | <5 |  |  | <20 |  | 0.4u |  |

*The total number of cancers reported here is slightly lower than the actual number in the dataset due to mismatched sex (see section 2.1). This figure also excludes non-melanoma skin cancers. Figures in bold are statistically significant at 5% level. U: unreliable age- sex- standardisation due to n<20 cases.

S4. All-cause cancer mortality (raw numbers [n], percentages [%], Crude Mortality Rate [CMR], Standardized Mortality Ratios [SMR] with 95% Confidence Intervals [CI]), for all-cause of death as cancer. CMR are reported per 100,000 person-years and SMR are age- sex- standardised rate ratios.

| Cancer category | Intellectual Disabilities (n=17,203) | | | General Population (n=566,061) | | | SMR  (95% CI) |
| --- | --- | --- | --- | --- | --- | --- | --- |
|  | n | % | CMR | n | % | CMR |  |
| Number of all-cause cancer deaths | 537 | 100·0% | 393·2 | 22,467 | 100·0% | 479·7 | **1·29 (1·17, 1·41)** |
| Digestive organs | 160 | 29·8% | 117·1 | 6,429 | 28·6% | 137·3 | **1·26 (1·06, 1·50)** |
| Colorectal | 70 | 13·0% | 51·3 | 2,694 | 12·0% | 57·5 | **1·37 (1·06, 1·78)** |
| Oesophageal | 25 | 4·7% | 18·3 | 898 | 4·0% | 19·2 | 1·20 (0·77, 1·86) |
| Stomach | 20 | 3·7% | 14·6u | 646 | 2·9% | 13·8 | 1·47u (0·92, 2·37)u |
| Liver | 14 | 2·6% | 10·3u | 729 | 3·2% | 15·6 | 1·04u (0·59, 1·85)u |
| Pancreas | <20 |  | 8·8u | 847 | 3·8% | 18·1 | 0·80u (0·43, 1·50)u |
| Respiratory & Intrathoracic organs | 79 | 14·7% | 57·8 | 5,880 | 26·2% | 125·6 | **0·74 (0·59, 0·94)** |
| Lung | 67 | 2·5% | 49·1 | 5,005 | 22·3% | 106·9 | **0·74 (0·57, 0·96)** |
| Unknown primary origin (metastases) | 57 | 10·6% | 41·7 | 1,284 | 5·7% | 27·4 | **2·53 (1·90, 3·38)** |
| Lymphoid, haematopoietic & related tissue | 46 | 8·6% | 33·7 | 1,824 | 8·1% | 39·0 | 1·23 (0·90, 1·69) |
| Hodgkin's & Non-Hodgkin's lymphoma | 24 | 4·5% | 17·6 | 674 | 3·0% | 14·4 | **1·72 (1·11, 2·66)** |
| Lymphoid leukaemia | <10 |  | 4·4u | 274 | 1·2% | 5·9 | 1·30u (0·54, 3·14)u |
| Myeloid leukaemia | 5 | 0·9% | 3·7u | 324 | 1·4% | 6·9 | 0·80u (0·29, 2·20)u |
| Leukaemia of unspecified cell type | <5 |  |  | 5 | 0·0% | 0·1u |  |
| Urinary tract | 44 | 8·2% | 32·2 | 1,580 | 7·0% | 33·7 | 1·37 (0·99, 1·89) |
| Kidney | 22 | 4·1% | 16·1 | 451 | 2·0% | 9·6 | **2·14 (1·36, 3·36)** |
| Bladder | 13 | 2·4% | 9·5u | 709 | 3·2% | 15·1 | 1·04u (0·58, 1·89)u |
| Central Nervous System | 15 | 2·8% | 11·0u | 492 | 2·2% | 10·5 | 1·08u (0·63, 1·87)u |
| Lip, oral cavity & pharynx | 13 | 2·4% | 9·5u | 581 | 2·6% | 12·4 | 1·04u (0·57, 1·88)u |
| Melanoma (skin) | 6 | 1·1% | 4·4u | 280 | 1·2% | 6·0 | 1·36u (0·57, 3·28)u |
| *Non-melanoma skin* | *36* | *0·2%* | *89·3* | *2,805* | *0·5%* | *257·3* | *0·50 (0·41, 0·61)* |
| Mesothelial and soft tissue | <5 |  |  | 422 | 1·9% | 9·0 |  |
| Thyroid & other endocrine glands | <5 |  |  | 73 | 0·3% | 1·6 |  |
| Thyroid | <5 |  |  | 39 | 0·2% | 0·8 |  |
| Bone and articular cartilage | <5 |  |  | 62 | 0·3% | 1·3 |  |

*The total number of cancers reported here is slightly lower than the actual number in the dataset due to mismatched sex (see section 2.1 for more detail). This figure also excludes non-melanoma skin cancers. Figures in bold are statistically significant at 5% level.

S5. All-cause cancer mortality by sex (raw numbers [n], percentages [%], Crude Mortality Rate [CMR], Standardized Mortality Ratios [SMR] with 95% Confidence Intervals [CI]), for all-cause of death as cancer. CMR are reported per 100,000 person-years and SMR are age- standardised rate ratios.

| Cancer category | Intellectual Disabilities (n=17,203) | | | | | | General Population sample (n=566,061) | | | | | | SMR (95% CI) | |
| --- | --- | --- | --- | --- | --- | --- | --- | --- | --- | --- | --- | --- | --- | --- |
|  | Females | | | Males | | | Females | | | Males | | |  |  |
|  | n | % | CMR | n | % | CMR | n | % | CMR | n | % | CMR | Females | Males FI |
| Number of all-cause cancer deaths | 263 | 100·0% | 438·3 | 274 | 100·0% | 357·8 | 10,860 | 100·0% | 438·9 | 11,607 | 100·0% | 525·4 | **1·44 (1·26, 1·64)** | **1·14 (1·00, 1·30)** |
| Digestive organs | 62 | 23·6% | 103·3 | 98 | 35·8% | 128·0 | 2,857 | 26·3% | 115·5 | 3,572 | 30·8% | 161·7 | **1·30 (1·00, 1·70)** | 1·23 (0·99, 1·54) |
| Colorectal | 27 | 10·3% | 45·0 | 43 | 15·7% | 56·2 | 1,258 | 11·6% | 50·9 | 1,436 | 12·4% | 65·0 | 1·35 (0·90, 2·03) | 1·39 (0·99, 1·94) |
| Oesophageal | 7 | 2·7% | 11·7u | 18 | 6·6% | 23·5u | 312 | 2·9% | 12·6 | 586 | 5·0% | 26·5 | 1·13 (0·52, 2·46)u | 1·24 (0·73, 2·10)u |
| Stomach | 8 | 3·0% | 13·3u | 12 | 4·4% | 15·7u | 259 | 2·4% | 10·5 | 387 | 3·3% | 17·5 | 1·59 (0·75, 3·36)u | 1·40 (0·76, 2·58)u |
| Liver | 7 | 2·7% | 11·7u | 7 | 2·6% | 9·1u | 271 | 2·5% | 11·0 | 458 | 3·9% | 20·7 | 1·64 (0·75, 3·61)u | 0·69 (0·30, 1·56)u |
| Pancreas | <5 |  |  | 8 | 2·9% | 10·5u | 430 | 4·0% | 17·4 | 417 | 3·6% | 18·9 |  | 1·02 (0·47, 2·21)u |
| Respiratory & Intrathoracic organs | 36 | 13·7% | 60·0 | 43 | 15·7% | 56·2 | 2,818 | 25·9% | 113·9 | 3,062 | 26·4% | 138·6 | 0·79 (0·56, 1·11) | 0·70 (0·51, 0·97) |
| Lung | 30 | 11·4% | 50·0 | 37 | 13·5% | 48·3 | 2,441 | 22·5% | 98·7 | 2,564 | 22·1% | 116·1 | 0·77 (0·53, 1·13) | 0·71 (0·50, 1·02) |
| Metastatic cancers of unknown primary origin | 27 | 10·3% | 45·0 | 30 | 10·9% | 39·2 | 716 | 6·6% | 28·9 | 568 | 4·9% | 25·7 | **2·50 (1·66, 3·76)** | **2·58 (1·73, 3·84)** |
| Haematopoietic | 19 | 7·2% | 31·7u | 27 | 9·9% | 35·3 | 785 | 7·2% | 31·7 | 1,039 | 9·0% | 47·0 | 1·32 (0·81, 2·14)u | 1·17 (0·77, 1·78) |
| Hodgkin's & Non-Hodgkin's lymphoma | 13 | 4·9% | 21·7u | 11 | 4·0% | 14·4u | 294 | 2·7% | 11·9 | 380 | 3·3% | 17·2 | **2·31 (1·29, 4·16)u** | 1·26 (0·65, 2·42)u |
| Lymphoid leukaemia | <5 |  |  | 5 | 1·8% | 6·5u | 104 | 1·0% | 4·2 | 170 | 1·5% | 7·7 |  | 1·88 (0·72, 4·91)u |
| Myeloid leukaemia | <5 |  |  | <5 |  |  | 159 | 1·5% | 6·4 | 165 | 1·4% | 7·5 |  |  |
| Leukaemia of unspecified cell type | <5 |  |  | <5 |  |  | <5 |  |  | <5 |  |  |  |  |
| Urinary tract | 17 | 6·5% | 28·3u | 27 | 9·9% | 35·3 | 591 | 5·4% | 23·9 | 989 | 8·5% | 44·8 | **1·78 (1·08, 2·94)u** | 1·12 (0·74, 1·70) |
| Kidney | 9 | 3·4% | 15·0u | 13 | 4·7% | 17·0u | 179 | 1·6% | 7·2 | 272 | 2·3% | 12·3 | **2·89 (1·46, 5·72)u** | 1·64 (0·92, 2·92)u |
| Bladder | 7 | 2·7% | 11·7u | 6 | 2·2% | 7·8u | 261 | 2··4% | 10·6 | 448 | 3·9% | 20·3 | 1·87 (0·85, 4·12)u | 0·56 (0·24, 1·30)u |
| Female genital organs | 44 | 16·8% | 73·3 |  |  |  | 1,089 | 10·0% | 44·0 |  |  |  | **2·22 (1·61, 3·08)** |  |
| Body of Uterus | 19 | 7·3% | 31·7u |  |  |  | 346 | 3·2% | 14·0 |  |  |  | **3·00 (1·84, 4·90)u** |  |
| Ovary | 18 | 6·9% | 30·0u |  |  |  | 374 | 3·4% | 15·1 |  |  |  | **2·75 (1·65, 4·60)u** |  |
| Vulva | <5 |  |  |  |  |  | 61 | 0·6% | 2·5 |  |  |  |  |  |
| Cervical | <5 |  |  |  |  |  | 98 | 0·9% | 4·0 |  |  |  |  |  |
| Breast | 43 | 16·4% | 71·7 |  |  |  | 1,247 | 11·5% | 50·4 |  |  |  | **2·07 (1·50, 2·87)** |  |
| Male genital organs |  |  |  | 37 | 13·6% | 48·3 |  |  |  | 1,957 | 16·9% | 88·7 |  | 1·29 (0·91, 1·83) |
| Prostate |  |  |  | 22 | 8·1% | 28·7 |  |  |  | 1,379 | 11·9% | 62·5 |  | 1·24 (0·80, 1·92) |
| Testicular |  |  |  | <5 |  |  |  |  |  | 15 | 0·1% | 0·7u |  |  |
| Central Nervous System | 9 | 3·4% | 15·0u | 6 | 2·2% | 7·8u | 209 | 1·9% | 8·5 | 283 | 2·4% | 12·8 | 1·73 (0·87, 3·44)u | 0·61 (0·26, 1·46)u |
| Lip, oral cavity & pharynx | 5 | 1·9% | 8·3u | 8 | 2·9% | 10·5u | 174 | 1·6% | 7·0 | 407 | 3·5% | 18·4 | 1·53 (0·62, 3·82)u | 0·82 (0·37, 1·81)u |
| Melanoma (skin) | <5 |  |  | <5 |  |  | 115 | 1·1% | 4·7 | 165 | 1·4% | 7·5 |  |  |
| *Non-melanoma skin* | *13* | *0·2%* | *81·7* | *23* | *0·2%* | *95·3* | *1,110* | *0·4%* | *213·1* | *1,695* | *0·6%* | *306·8* | ***0·53 (0·39, 0·72)u*** | ***0·47 (0·37, 0·61)*** |
| Mesothelial and soft tissue | <5 |  |  | <5 |  |  | 161 | 1·5% | 6·5 | 261 | 2·2% | 11·8 |  |  |
| Thyroid & other endocrine glands | <5 |  |  | <5 |  |  | 46 | 0·4% | 1·9 | 27 | 0·2% | 1·2 |  |  |
| Thyroid | <5 |  |  | <5 |  |  | 27 | 0·2% | 1·1 | 12 | 0·1% | 0·5u |  |  |
| Bone and articular cartilage | <5 |  |  | <5 |  |  | 28 | 0·3% | 1·1 | 34 | 0·3% | 1·5 |  |  |

Note: non-melanoma skin cancers were not included in the rates calculations in the denominator. Figures in bold are statistically significant at 5% level.

U: unreliable age- sex- standardisation due to n<20 cases.
